# Ketamine for the multivariate effect of PTSD: Systematic review and meta-analysis

**DOI:** 10.1101/2021.06.13.21258836

**Authors:** Rui Du, Kun Niu, Guofang Lu, Yulong Shang

## Abstract

The aim of this systematic review and meta-analysis was to examine the efficacy, anti-effect of ketamine (intervention) for post-traumatic stress disorder (PTSD) patients during analgesia proceeding and mental illness treatment methods, in comparison with control (midazolam, opioid, saline or placebo). The bibliographic databases Cochrane, Embase, Pubmed and Web of Science were searched from inception to 23 May 2021 for randomized controlled trials, case-control and cohort studies included. For continuous and dichotomous outcomes, respectively, we calculated the mean difference using the inverse-variance method and the risk ratio with the Mantel–Haenszel method. In all, ten trials with 705 patients were included. Confirmed by meta-analysis, ketamine didn’t increase the prevalence of PTSD by a risk ratio (95 % CI) of 0.86 (0.61 to 1.20), p = 0.38 in 3 trials with 503 patients. Evidence of a difference was found in the PTSD-scales taken between ketamine and control during short durations (months), with a mean difference (95 % CI) of 2.45 (1.33 to 3.58), p < 0.001 in three trials with 65 patients. Another evidence is shown in chronic PTSD (years), with a mean difference (95 % CI) of −3.66 (−7.05 to −0.27), p = 0.03 in three trials with 91 patients. Sub-group analysis underlined the increased benefit of ketamine administration for those in whom the procedure was more than one week in the chronic PTSD group. The adoption of ketamine for the short duration of PTSD is in avoidance, but for chronic PTSD is recommended and, in the opinion of the authors, should be considered as a new therapy in view of its potential to ameliorate arousal, avoidance and dissociative symptoms, neuroticism after trauma needing more animal research and clinical trials.

## Introduction

According to the fifth updated diagnostic criteria for post-traumatic stress disorder (PTSD), it belongs to “Trauma- and Stressor-Related Dis-order” [1]. Patients with PTSD have pain, repeated recall of the traumatic experience, avoiding traumatic situations, depression, anxiety, substance-use disorders and other symptoms [2]. And PTSD is the most prevalent psychopathological consequence of exposure to traumatic events [3]. The lifetime prevalence of PTSD ranging from 1.3 to 12.2%, and the 1-year prevalence is 0.2 to 3.8 %; the varies according to social background and country of residence [4]. PTSD is also associated with suicidal behavior [5], but the relationship is still unclear. PTSD is currently no specific treatment; existing treatments include psychotherapy, medication and innovative interventions, etc. [6-9]. Due to multiplicity and interdependence of biologic features [10-15] and complicated pathogenesis, the pressing need for diagnostic, prognostic, and therapeutic biomarkers about the pathogenesis of PTSD and treatment targets.

Ketamine acts as blocking the excitatory receptor antagonist N-methyl-d-aspartate receptor (NMDA) [16,17] is a versatile agent primarily utilized as a dissociative anesthetic anesthetic [18-21], analgesic [22,23], anti-inflammatory [24,25], and antidepressant [26] drug has reignited research to understand the neurobiological mechanisms underlying these effects of ketamine. However, the molecular mechanisms involved in clinical therapeutic effects the action of ketamine is still unclear. In recent years, data suggest that intravenous ketamine (a single or repeated dose) can quickly reduce the severity of PTSD symptoms in chronic cases (14.6 (25.9), years (SD)) [27,28]. But the therapeutic effect of ketamine on PTSD needs further exploration.

In the present study, we conducted a systematic review and meta-analysis to evaluate the relationship between ketamine and PTSD. We did not detect any discernible effect of ketamine on the development of PTSD. Patients with chronic PTSD exhibited significant improvements than acute or short-term PTSD patients after ketamine treatment.

## Methods

This systematic review and network meta-analysis were registered with PROSPERO and follow the preferred reporting project for systematic reviews and meta-analysis (PRISMA) recommendations [29].

A detailed systematic review of the following databases was performed: Cochrane Central Register of Controlled Trials; Embase; Pubmed and Web of Science from inception to 23 May 2021. The following search terms were used for Pubmed: (‘ketamine’ OR ‘ketalar’ OR ‘calipsol’ OR ‘kalipsol’) AND (‘PTSD’ OR ‘posttraumatic stress disorder’ OR ‘posttraumatic stress symptoms’ OR ‘acute stress disorder’). Differently, RCT (Randomized Controlled Trial), Case-control study and Cohort study were included. Both dichotomous and continuous variables were separately included. Research articles were also extended to pre-published in BioRivx.

A preliminary screening of the abstract, if found to be suitable, the full text of this article would be further studied. Two authors (Rui and Guofang) independently reviewed all search results based on inclusion and exclusion criteria. The divergent research items were determined by independent researchers (Kun). All included trials were reviewed by authors to ensure that they met the eligibility criteria. The risk of bias was used by Review Manager 5.3. This included: random sequence generation, allocation concealment, blinding of participants and personnel, blinding of outcome assessment, incomplete outcome data, selective reporting and other sources of bias. Disagreements in the process were resolved through discussion until a consensus was reached.

Results related to the terms of ‘ketamine’ and ‘PTSD’ were included referred to RCT, Case-control and Cohort studies. We excluded: correspondence, poster abstract, no results of NCT (National ClinicalTrial.gov, paired comparison and unable to obtain the full text.

Two investigators (Rui and Guofang) independently screened the literature based on the above inclusion and exclusion criteria. By revisiting the original text and performing any appropriate calculations or conversions, any discrepancies found would be resolved. Intuitively extracted data includes: the first author, publication time, participants, study interventions, types of outcomes, reasons for exclusion, study control and duration of follow-up.

The primary outcomes were giving ketamine analgesia during treatment on the battlefield to the risk of PTSD in the later stage of the wounded or burn patients. The second outcomes were the rate of the effect of ketamine on the symptom performance of wounded patients with confirmed PTSD (classified as early, usually several months), based on multiple psychological assessment scales. The third outcomes were related to the long duration (several years) of PTSD compared to the second outcome. The Mantel-Haenszel (M-H) method was used to pool dichotomous data and to compute the risk ratio (RR) with its corresponding 95 % confidence interval (CI) was calculated. The inverse variance (IV) method was used to pool continuous data and to calculate standard mean difference with 95 % CI. Random effects models were used with or without apparent heterogeneity based on I^2^ > 50 % compared to the fixed effects model (I^2^ < 50 %) [30,31]. Sensitivity analysis was performed to assess the stability of the meta-analysis. P < 0.05 was considered statistically significant.

The software used was Review Manager (RevMan, version5.3.5. Copenhagen: The Nordic Cochrane Centre, The Cochrane Collaboration, 2014).

## Results

The process of retrieving and screening the research was shown in a flowchart in Fig. 1. Iinitial searching found 3252 articles, including 328 duplicates, leaving 2924 to be screened according to the previously formulated retrieval strategies. After exclusion by title or abstract of 2901 articles, 23 full text articles were reviewed and assessed for eligibility. Of these, 13 were further excluded because 2 were correspondence without chart data [32,33], 2 were poster abstract without chart data [34,35], 2 were NCT finished without results posted [NCT02398136, NCT02655692], 3 were abstracts unavailable to obtain full texts [36-38] and 4 were paired comparison unconsidered to include in Meta-analysis (Fig.1) [39-42]. Finally, 10 studies that met (5 RCT studies, 3 Case-control studies and 2 Cohort studies) our predetermined inclusion criteria and were incorporated in this study. Details of the screening process were shown in Fig. 1 and the results of the risk of bias assessment were found in Fig. 2. All 10 studies reported the effect of ketamine analgesia on the early occurrence of PTSD after battlefield rescue or burned patients [43-45], symptom changes in short duration (months) of PTSD patients with a dose of ketamine (0.5 mg/kg) [27,28,46] and the same for long duration (years) of PTSD patients [47-50].

**Figure 1.**
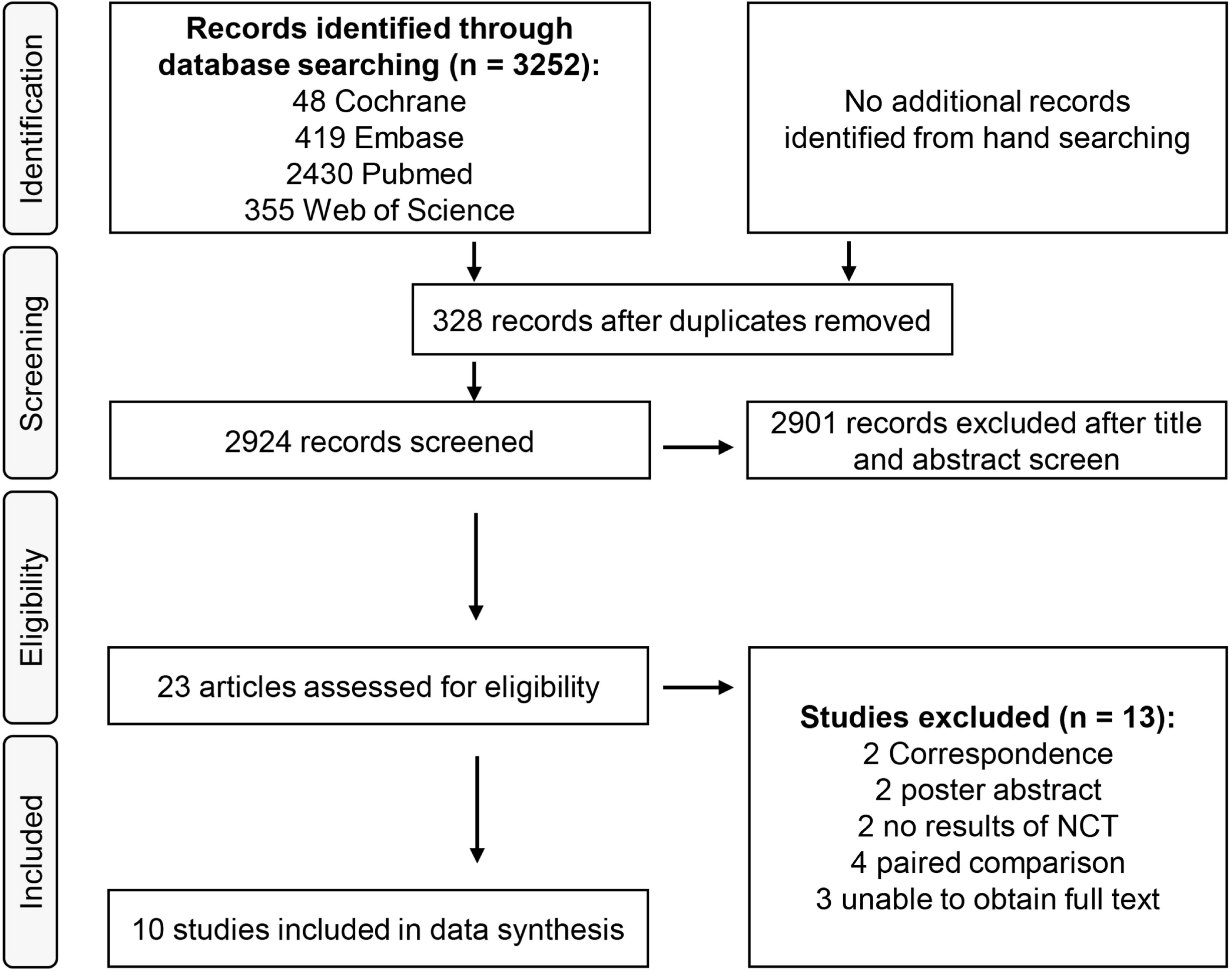
PRISMA flowchart of included and excluded studies.

**Figure 2.**
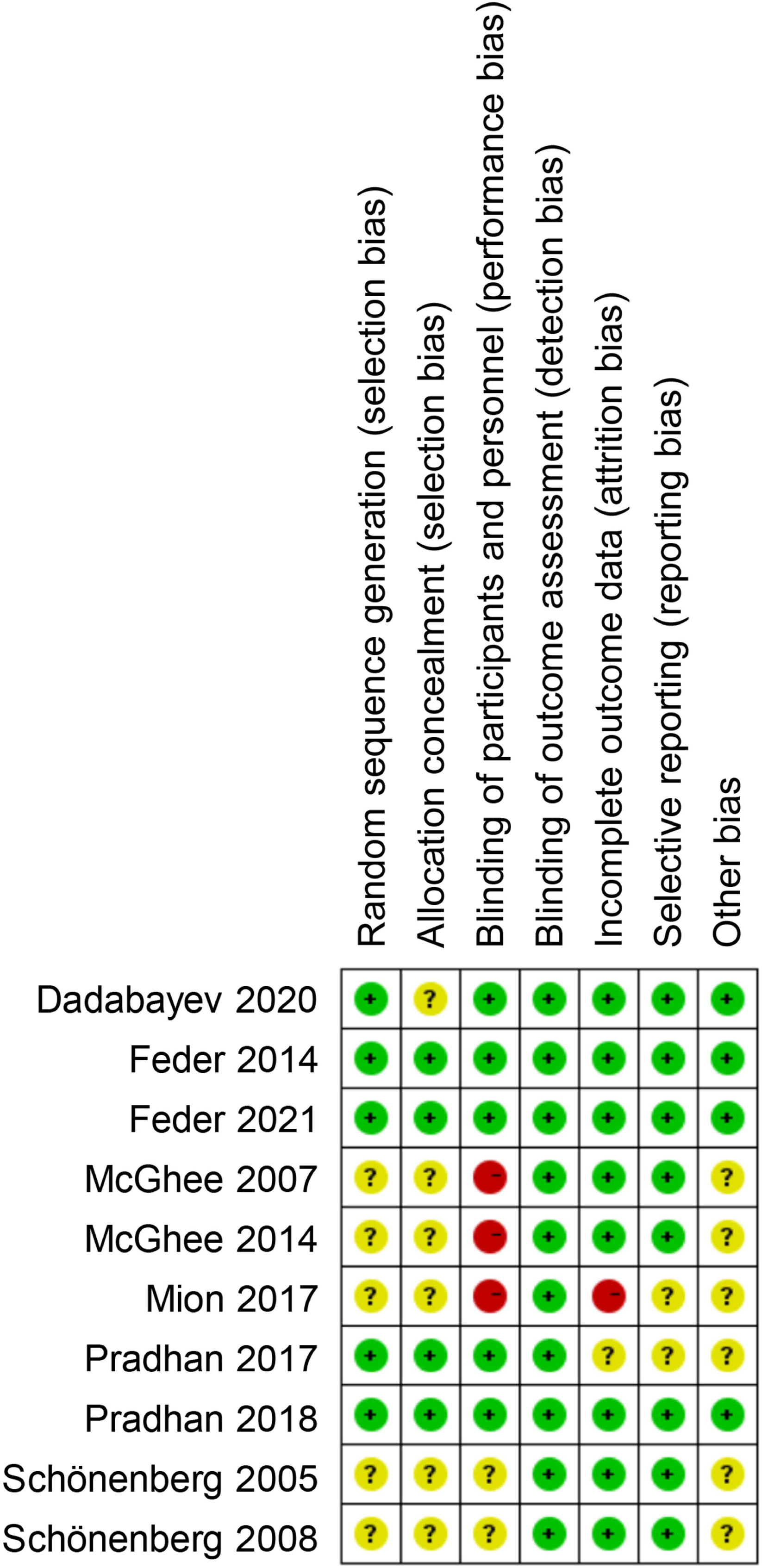
Summary table illustrating risk of bias results for included studies. Green circle, low risk; yellow circle, some concerns; red circle, high risk.

The 10 studies involved a total of 705 patients, and were allocated to eligible study groups (Fig. 1). Characteristics of the trials were presented in Table 1. Of these, a total of 442 patients were assigned to ketamine or (S)-ketamine and 263 were assigned to control (i.e. ketorolac, midazolam, normal saline, opioid or placebo, Table 1). Patients in 5 studies (Table 1) [43-46,48] received diagnosis of PTSD assessed with a score of PTSD Checklist (PCL) [51]. Patients in 4 studies (Table 1) [27,28,46,48] received the Clinician-Administered PTSD Scale (CAPS) [52]. Patients in 4 studies (Table 1) [27,28,47,49] received Impact of Event Scale-Revised (IES-R) [53]. Patients in two studies (Table 1) [49,50] received Acute Stress Disorder Scale (ASDS) [54].

**Table 1.**
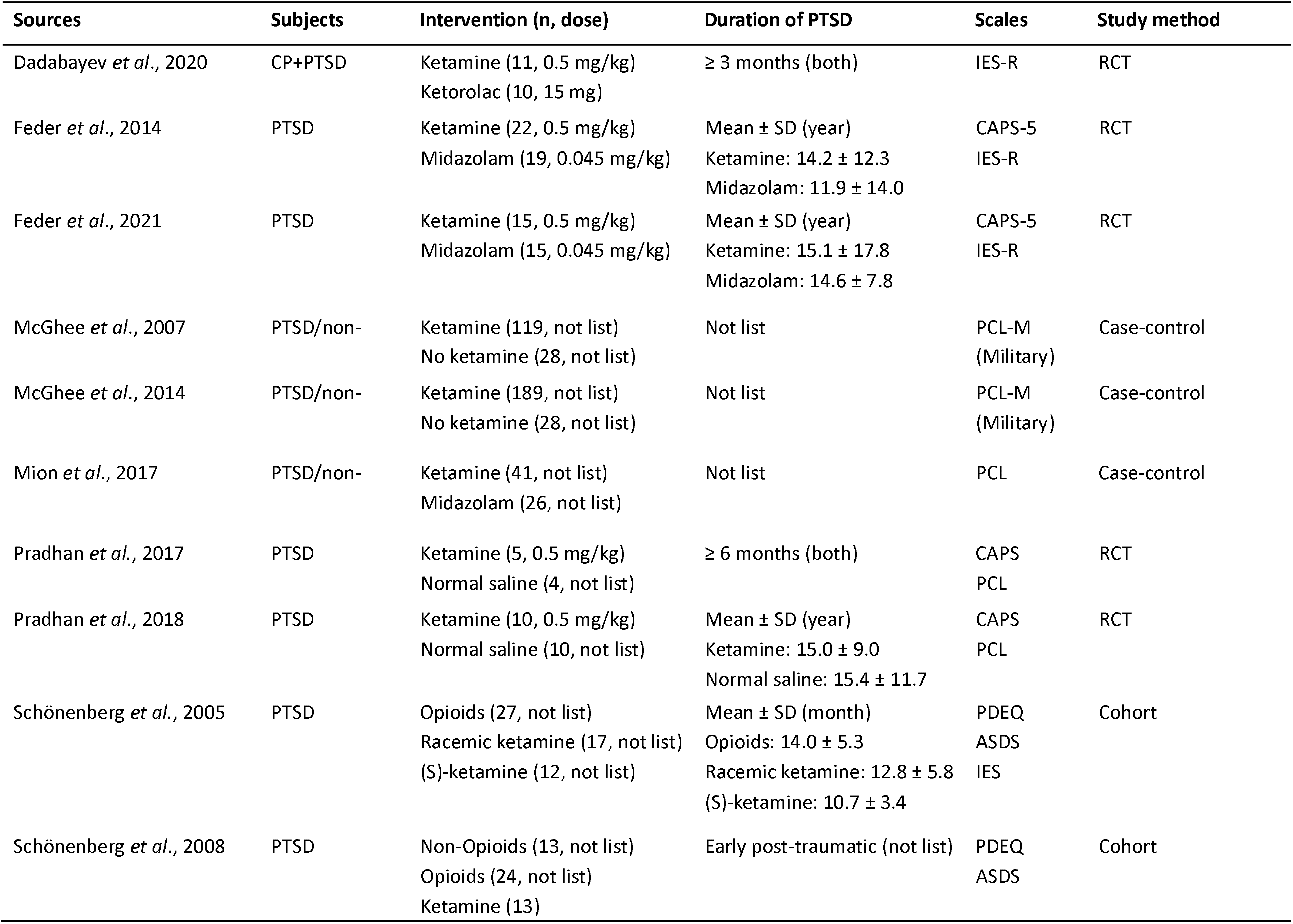
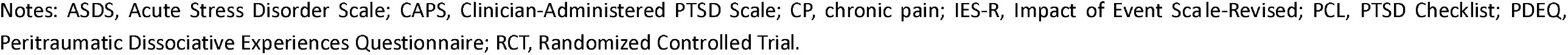
Included study characteristics, intervention, duration, scales and study method.

Three studies [43-45] including 503 patients (ketamine 349, control 154) in battlefield reported no significant reduction of PTSD prevalence (Fig. 3) with ketamine (no dose list) by a risk ratio (95 % CI) of 0.86 (0.61-1.20), p = 0.38, I2 = 52 % compared with control.

**Figure 3.**
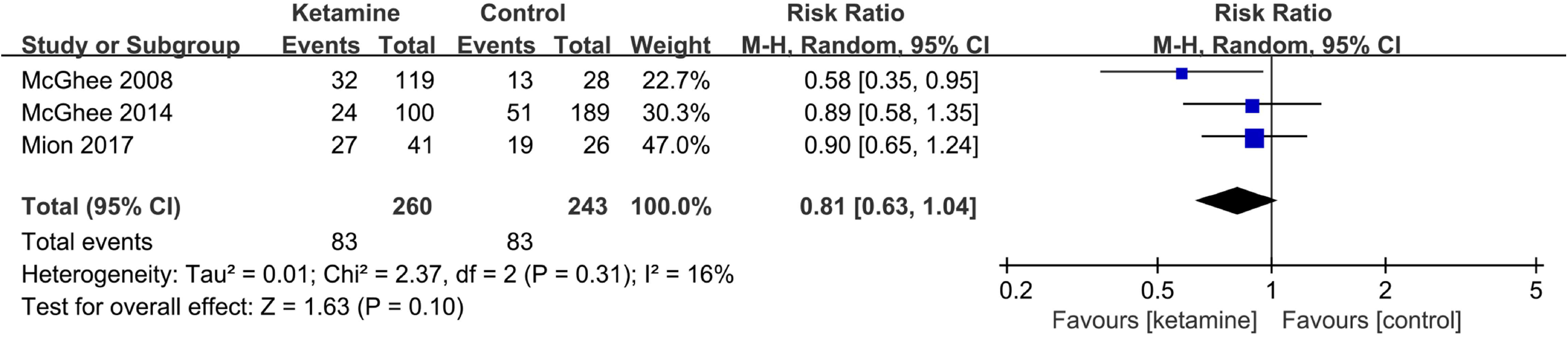
Forest plot of the prevalence of PTSD in battlefield analgesia by ketamine. M–H, Mantel–Haenszel.

Three studies [47,48,50] including 67 patients (ketamine 29, control 38) reported scores changed in short duration of PTSD (months) by scales of ASDS (four subscales, including dissociation, reexperiencing, avoidance and hyperarousal) [50], IES-R, PCL and CAPS (Fig. 4). Compared with control, ketamine (0.5 mg/kg) aggravated PTSD symptoms (Fig. 4) by a mean difference (95 % CI) of 2.45 (1.33-3.58), p < 0.001, I2 = 0%. Varying numbers of patients were analyzed for each time-point (1 day: ketamine 16, control 14; < 3 days: ketamine 13, control 24; 1 week: ketamine 11, control 10). Subgroup analysis showed no influence of ketamine post-infusion during different times on this outcome (p = 0.16). Including one study without infusion time [49], ketamine aggravated PTSD symptoms (Fig. S1) by a mean difference (95 % CI) of 1.72 (0.95-2.48), p < 0.001, I2 = 0 %.

**Figure 4.**
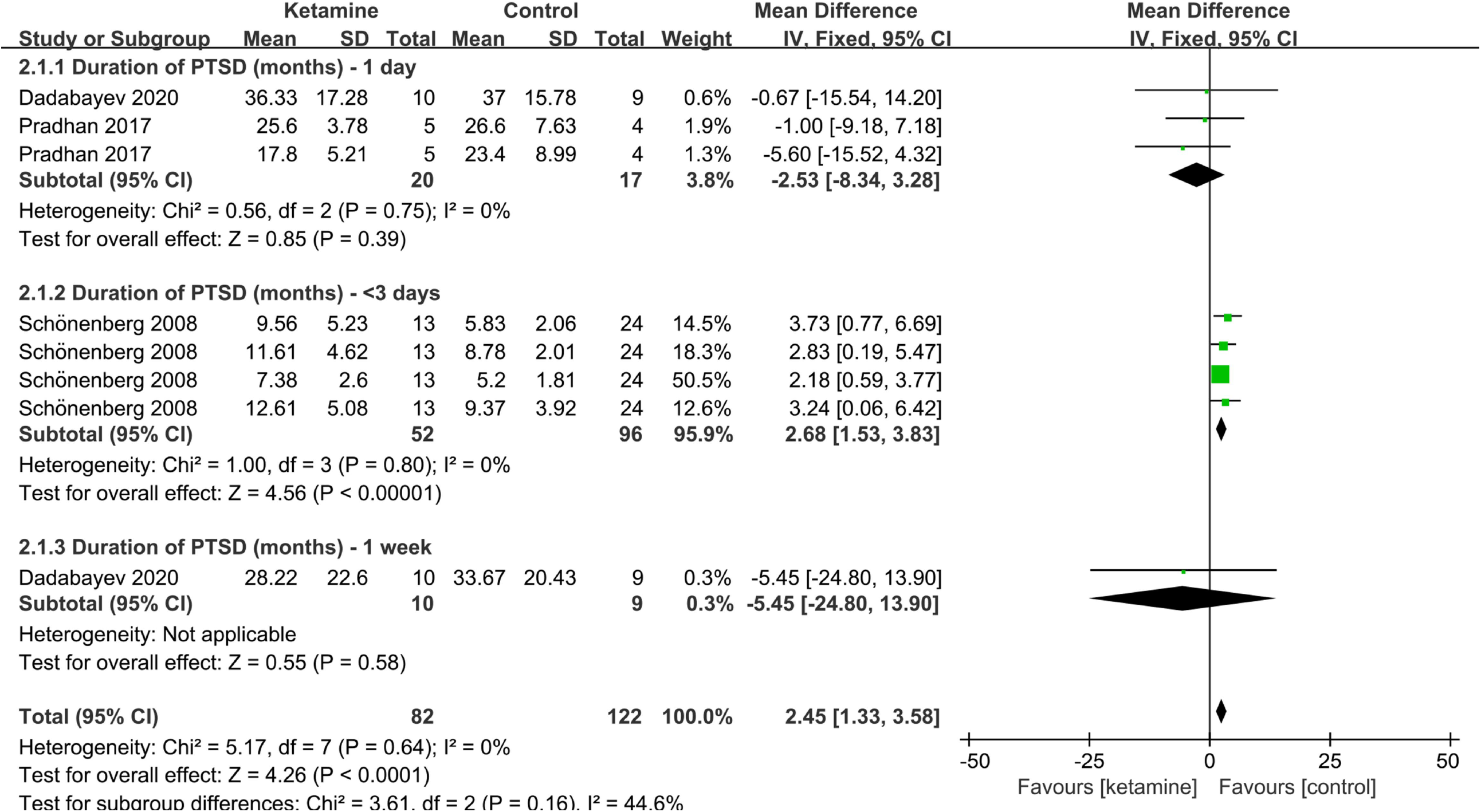
PTSD-scale scores of short duration (months) by ketamine administration (ketamine vs. control). IV, inverse variance; SD, standard deviation.

Our third co-primary outcome, including 91 patients (ketamine 47, control 44) [27,28,46] reported scores changed in long duration of PTSD (years) by scales of CAPS, IES-R and PCL (Fig. 5). Compared with control, ketamine (0.5 mg/kg) relieved PTSD symptoms (Fig. 5) by a mean difference (95 % CI) of −3.66 (−7.05--0.27), p = 0.03, I2 = 35%. Varying numbers of patients were analyzed for each time-point (1 day: ketamine 25, control 25; 1 week: ketamine 22, control 19; 2 weeks: ketamine 15, control 15). Subgroup analysis showed the long-term influence of decreasing assessment scores (relief effect) after infusion (p = 0.007). Mean difference (95 % CI) for 1 day, 1 week and 2 weeks were respectively, −0.19 (−3.34-2.96), −11.02 (−19.61--2.43) and −8.55 (−14.41--2.69).

**Figure 5.**
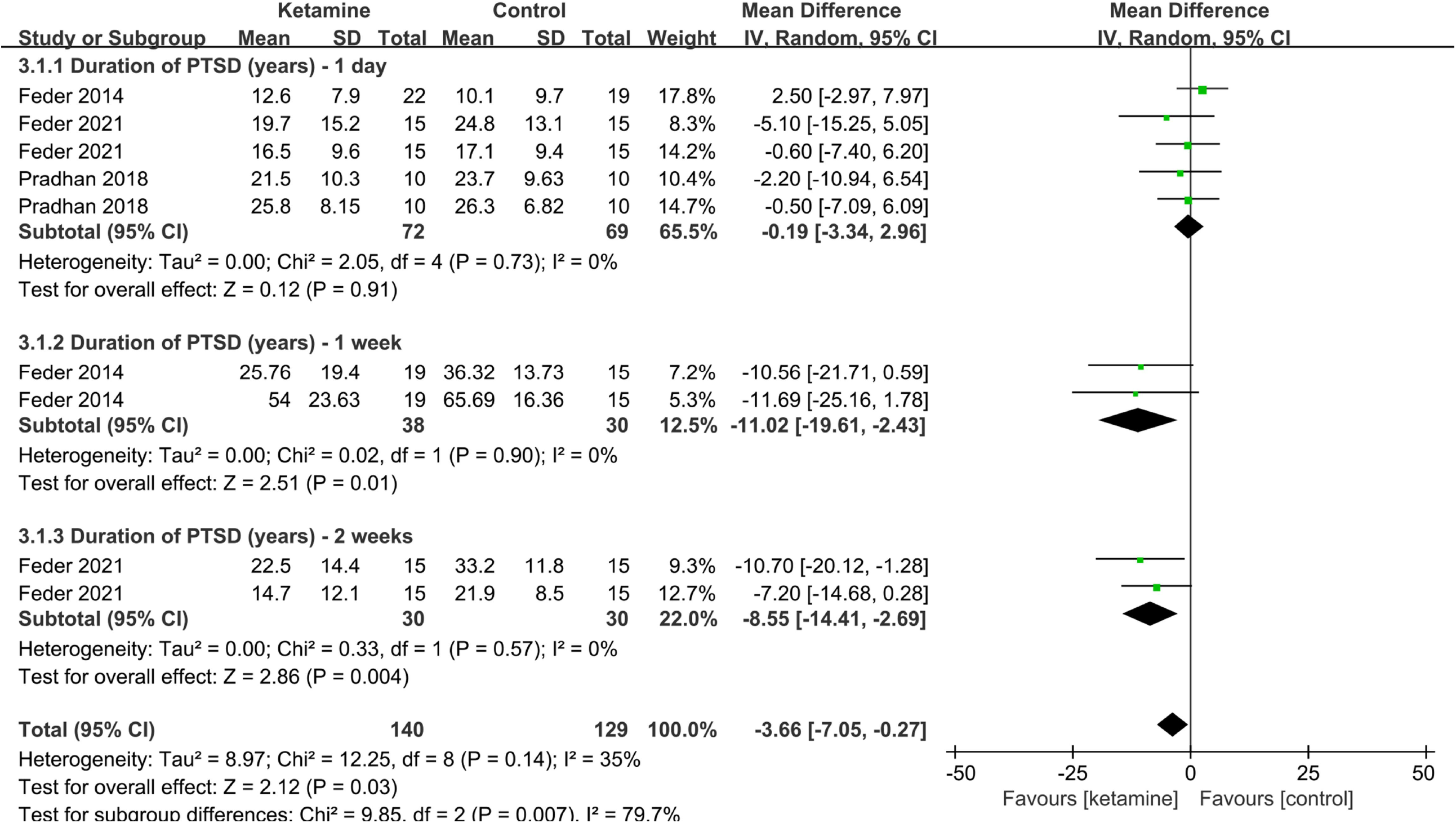
PTSD-scale scores of long duration (chronic, years) patients by ketamine. administration (ketamine vs. control). IV, inverse variance; SD, standard deviation.

## Discussion

In the present study, we identified that there was no effect on the development of PTSD for injured soldiers (e.g. firearm, explosive, burn or accident) on battlefield who was receiving ketamine or not. The symptoms were aggravated for the patients who had been diagnosed of PTSD with a short duration of disease (months) after the administration of ketamine. However, the symptoms were ameliorated for the patients with a chronic PTSD (years) sfter ketamine treatment. The results indicate that ketamine treatment was not related to the development of PTSD but was associated with the disease duration in PTSD patients.

It is traditionally believed that ketamine administration is associated with an increased incidence of PTSD. However, in our study, for injury soldiers, there is no significant difference between the development of PTSD and the treatment of ketamine. Although it is not clear why there is a difference in results, our findings are also supported by Mion et al [45] in that ketamine did not increase the high risk of PTSD. Administration of ketamine during a stressful event may increase the prevalence, reduce the preventive effect, or have no effect on the subsequent development of PTSD [43,45].

To our surprise, we found that ketamine can significantly alleviate the symptoms of PTSD with a long-term course, and without obvious symptoms of psychosis or mania. This has application significance for the clinical therapeutic effect of ketamine. Previous reports by Kyle et al. who used ketamine on Major Depressive Disorder (MDD), and found that rapid antidepressant effects of intranasal ketamine on MDD [26,55]. Interestingly, there has a similar therapeutic effect on PTSD after ibuprofen, a NSAIDs [56]. Possible mechanisms underlying ketamine’s therapeutic effects could be related to ketamine could rapidly increase synaptic connections in the prefrontal cortex and reverse the behavioral and neuronal changes caused by chronic stress in rats, partly by activating mammalian-targeted rapamycin signaling pathways and stimulation of brain-derived neurotrophic factor signals [27,57,58]. In addition, repeated injections of ketamine are safe for patients with chronic PTSD and are generally well tolerated, with only short-term mental and hemodynamic side effects [28].

On the contrary, through comprehensive data analysis, we also found that ketamine can aggravate the symptoms for diagnoses of PTSD with a short-term course, including within a week. It is known that the half-life of ketamine under anesthesia is 2-3 hours, the psycho-simulating symptoms last up to 3 days [49], and the effect can be observed up to 1 week under a single dose (Fig. 4). One possibility is that ketamine overstimulates the stress-induced glutamate-glucocorticoid interaction in the early stage of trauma, which in turn leads to stronger dissociation symptoms and fragmentation to consolidate traumatic memories and aggravate the symptoms [49]. In addition, ketamine could rapidly induce synapses in the brain-derived neurotrophic factor (BDNF) pathway and increase pro-inflammatory cytokines, and then activates microglia to aggravate the symptoms of PTSD [59].

In summary, results from the current study demonstrated that in the analgesia proceeding, the development of PTSD will not be affected by receiving ketamine or not. For chronic course, but not acute or short-term course of disease, facilitated damage caused by PTSD after ketamine treatment.

## Limitation

We cannot exclude the possibility that the difference of ketamine’s effect is caused by different statistical methods and standards. In addition, the results confirm that treatment of ketamine could ameliorate the symptoms caused by PTSD. However, we need to further verify this at both the animal research level and clinical studies. And the definitely mechanism of the positive or negative effect of ketamine is still unclear.

## Data Availability

No data, models, or code were generated or used during the study (e.g., opinion or dataless paper).

## Acknowledgements

This work was supported by grants from National Natural Science Foundation of China (81773072).

## Author Contributions

Rui Du designed the study, searched articles and analyzed data. Kun Niu analysed data. Guofang Lu searched articles and wrote the manuscript. Yulong Shang provided academic advice to the study and revised the manuscript.

## Figure legends

**Figure S1.**
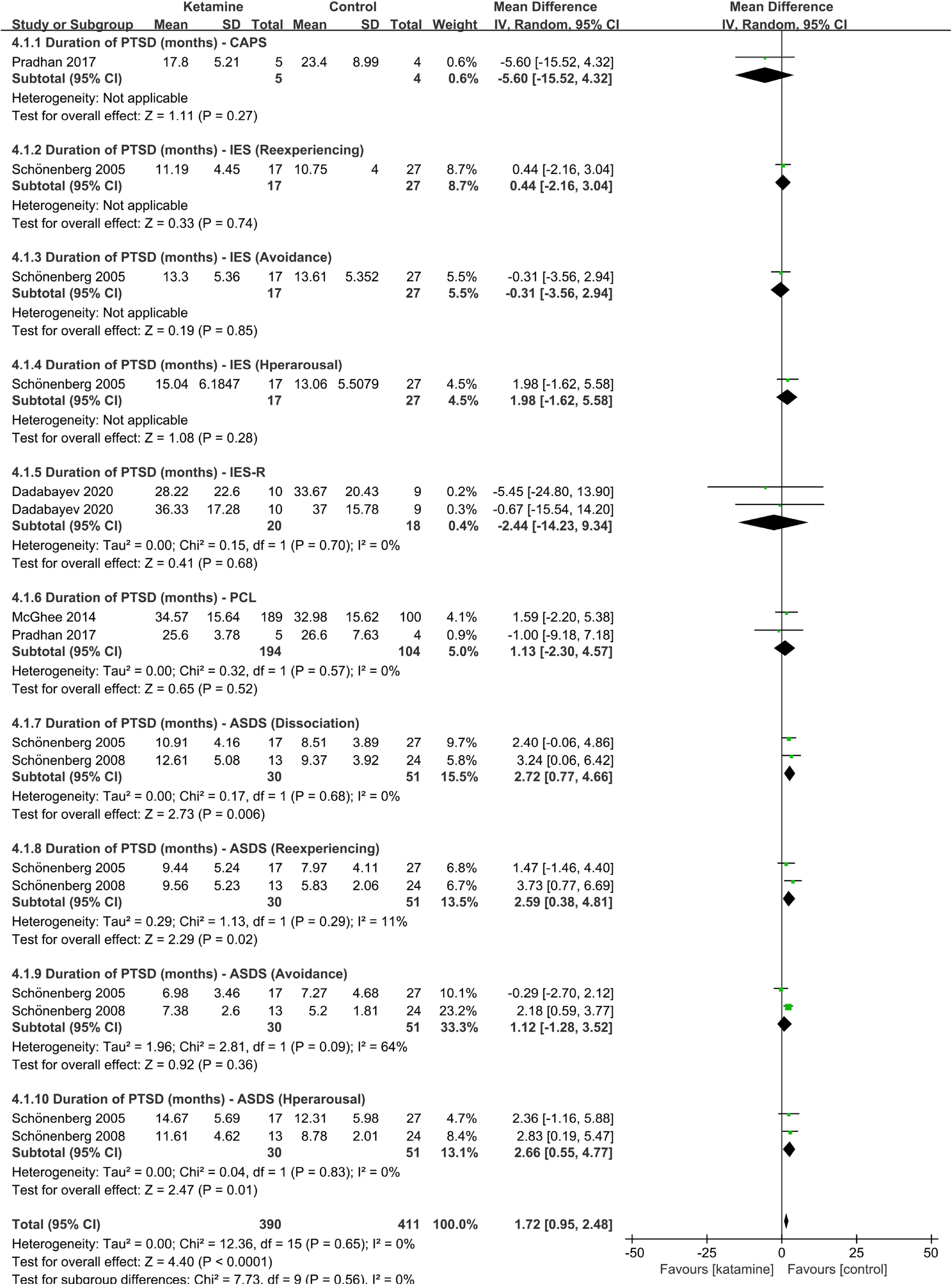
PTSD-scale scores of short duration (months) by ketamine administration. (ketamine vs. control, full scales). IV, inverse variance; SD, standard deviation.

